# Genetic associations of externalising and internalising symptoms with brain imaging and cell types among autistic individuals and the general population

**DOI:** 10.64898/2025.12.18.25342455

**Authors:** Adeniran Okewole, Yuanjun Gu, Amir Ebneabbasi, Marcin A. Radecki, Laramie Duncan, Hadar Segal-Gavish, Gizem Hazal Senturk, Nicholas Bray, Simon Braschi, Eden Hymanson, Holan Liang, Jennifer Parker, Vincent-Raphael Bourque, Sebastien Jacquemont, Taylor Thomas, Elise Robinson, Richard Bethlehem, Dennis van t Ent, Varun Warrier, Simon Baron-Cohen

## Abstract

Externalising and internalising symptoms span multiple psychiatric diagnoses. Although similar measures assess these traits in autistic and non-autistic populations, it remains unclear whether their polygenic influences and biological mechanisms align. This study compared genetic contributions to these symptoms in autistic individuals (SPARK, N=3,486) and the general population (ABCD, N=4,637; external datasets: Neff=523,150 externalising; Neff=132,260 internalising). Regression models tested associations between polygenic scores, demographics, and symptom outcomes. Genetic correlations were computed with 12 global and 2,159 regional brain phenotypes, and with 461 cell types across 31 superclusters. In both cohorts, higher symptoms correlated with lower maternal education, lower household income and polygenic scores for depression. The strongest associations were observed for externalising symptoms in the general population, showing negative correlations with cortical expansion and enrichment in hypothalamic and histaminergic neurons. These findings suggest shared genetic architectures but different neurobiological correlates of externalising and internalising symptoms across autism and the general population.

## MAIN

Beyond categorical diagnoses of mental health conditions, there is increasing awareness of a need for dimensional, transdiagnostic measures[1]. A common approach is to measure internalising (e.g., depression, anxiety, withdrawal, or somatic complaints) and externalising (e.g., impulsive, aggressive, or rule-breaking behavior) symptoms, which represent two different but correlated domains. Although these two domains are moderately correlated (or co-occurring), with phenotype-based r = 0.45 to 0.54[2], there is evidence pointing towards partly different genetic aetiologies, as also reflected in the broad factors observed in the genetics of psychiatric diagnoses[3]. These domains are also associated with different cognitive problems in childhood - externalising symptoms have been associated with problems with attention and executive functioning, while internalising symptoms are associated with problems in verbal fluency and memory[4]. The resulting academic, occupational and social challenges explain why externalising and internalising symptoms are associated with poor outcomes across the lifespan[5].

Externalising and internalising behaviours occur commonly in autistic persons as well as the general population, with the same diagnostic and therapeutic measures often used across populations. However, it is unclear if similar genetic and biological mechanisms underlie the two domains in the general population and among autistic individuals. An important source of understanding is the genetic contribution, regarding which still remains to be determined, as genetic correlation studies show high genetic correlation of autism with other mental and neurodevelopmental conditions when compared with the general population, but also substantial heterogeneity within the autism spectrum [6,7]. Dissecting the contribution of genetics and environmental factors however still remains challenging, as several models focus exclusively on one or the other.

Complementary with genetics, another key source of biological insight is neuroimaging. Measures of cortical macrostructure (such as surface area, gray matter volume, cortical thickness, folding index, intrinsic curvature index, local gyrification index, mean curvature and Gaussian curvature) have been found to be heritable, as have measures of cortical microstructure (such as fractional anisotropy, mean density, isotropic volume fraction, intracellular volume fraction or neurite density index, and orientation diffusion index), which are informative about cortical myelination, lamination and tissue properties such as density of axons and dendrites[8–11]. These brain phenotypes, measured using structural or diffusion-weighted magnetic resonance imaging (MRI), demonstrate variation in the general population, and have been shown to vary by brain region and to be associated with specific brain-related conditions such as depression, schizophrenia and autism[8,10,11]. With reference to internalising and externalising symptoms, Whittle et al [12] found that internalising and externalising symptoms were associated with different trajectories of cortical development during late childhood, with reduced thinning of the orbitofrontal cortex and postcentral gyrus respectively. A subsequent study[13] reported that externalising symptoms were predicted by larger and smaller subcortical gray matter volume in older (12 years) and younger (9 years) children respectively, again suggesting a relationship with developmental trajectory. The authors reported no predictive effects for internalising symptoms. This theme was also investigated by Nakua et al[5], who studied externalising symptoms among participants enrolled in the ABCD study. The authors found that reduction in externalising symptoms over a two-year period was predicted by higher baseline cortical thickness in the left pars triangularis and rostral middle frontal gyrus. There is however a paucity of studies which specifically included autistic individuals.

Imaging genetics represents a powerful combination of the tools of genetics and neuroimaging to better understand how genetic factors influence brain phenotypes. In addition, beyond localising brain regions via imaging, fine-grained analysis of specific cell types holds the promise of refining biological understanding and specifying targets for intervention. By combining genomic data of psychiatric conditions with human brain cell types, Yao et al [14] reported a convergence with functional connectivity. Their most indicative findings were however with respect to schizophrenia. An example of this approach combining brain imaging with cellular data in studying externalising and internalising symptoms is Davis et al [15,16], who found correlated but distinct genetic liability between internalising and externalising factors derived from general population datasets, using an integrated framework based on the Hierarchical Taxonomy of Psychopathology (HiTOP) and Research Domain Criteria (RDoC) frameworks[17,18], with specific imaging phenotypes and cell types implicated[15,16]. The HiTOP and RDoC frameworks represent a transdiagnostic challenge to the prevailing orthodoxy of categorical psychiatric diagnoses, which are oft-criticised for not sufficiently ‘carving nature at its joints’ in a way that yields unique biological insights and is therapeutically actionable. Using genomic structural equation modelling within these frameworks, Davis et al[15,16] found a best-fitting two-factor correlated model, representing externalising (EXT) and internalising (INT) spectra, with multivariate GWAS identifying 409 and 85 lead genetic variants for the EXT and INT spectra respectively. Using BrainXcan[19], the authors linked the genetic data to 327 imaging-derived phenotypes (IDPs) from structural and diffusion-weighted MRI. They found that 20 brain IDPs were associated with EXT, while INT was significantly associated with lower gray matter volume in the subcallosal cortex. Further, EXT was associated with 454 genes, with expression of EXT genes enriched in GABAergic, cortical and hippocampal neurons, while INT genes aligned more with GABAergic neurons. Taken together, these findings suggest a convergence of evidence that localises internalising and externalising symptoms to specific brain regions and pinpoints specific cell types that might be implicated.

However, the extent to which these findings apply to autistic individuals remains unclear. The genetic underpinnings of the relationship (if any) between externalising and internalising symptoms and brain structure also remains unclear. In this study, we aimed to compare the heritability of externalising and internalising symptoms in autistic individuals and the general population, and we questioned whether the common genetic variant profile of these symptom groups was comparable between autistic individuals and the general population. We also aimed to compare models of externalising and internalising symptoms with and without genetic data among autistic individuals, and asked whether polygenic profiles are informative for these symptom groups specifically in autistic individuals. Finally, we aimed to explore brain imaging and cellular enrichment of these symptom groups in autistic individuals and the general population, and we investigated whether there are distinct associations of the symptom groups with brain imaging phenotypes and cell types across the two populations.

## RESULTS

### Participant Characteristics

Participants in the SPARK dataset had a median registration age of 7 years (IQR=5 years), including 21.4% females and 15.2% with cognitive impairment. The median t scores on internalising and externalising symptoms was 65 (IQR= 13) and 59 (IQR= 16) respectively. Participants in the ABCD dataset had a median interview age of 119 months (IQR=14 months), including 47.3% females and 2.3% with cognitive impairment. The median t scores on internalising and externalising symptoms was 48 (IQR= 13) and 44 (IQR= 17) respectively. The composition of the other two general population datasets is described elsewhere[20,21].

### Heritabilities, Genetic Correlations and GWAS

To understand the polygenic architecture of externalising and internalising in the general population and autistic cohorts, we first investigated the SNP-based heritabilities of and genetic correlations among the two traits in different populations. SNP heritability for externalising symptoms in SPARK and ABCD was 0.26 (s.e.=0.11) and 0.22 (s.e.=0.08) respectively, while SNP heritability for internalising symptoms in SPARK and ABCD was 0.11 (s.e.=0.11) and 0.06 (s.e.=0.08) respectively (Supplementary Table ST1). Genetic correlation (rG) for externalising between SPARK and ABCD was 0.67 (s.e.=0.41); between SPARK externalising and Baselmans et al.[21] externalising was 0.31 (s.e.=0.10), and between ABCD externalising and Baselmans et al.[21] externalising was 0.75 (s.e.=0.11). Notably, the genetic correlations between externalising in the autistic cohort and the two external general population cohorts were moderate, suggesting differing polygenic correlates of externalising in the autistic population. LDSC did not output rG for internalising symptoms, possibly due to low and non-significant SNP heritability. There were no genome-wide significant loci for any of the GWAS conducted (see Supplementary Figure 1).

### Regression Models

In order to investigate the contribution of polygenic variation and environmental variables to internalising and externalising symptoms in autistic individuals and the general population, we computed regression models for SPARK and ABCD. Across the two sets of models (without and with polygenic scores, for SPARK and ABCD), presence of cognitive impairment was significantly associated with higher externalising scores (Figure 1 and Supplementary Table ST2). Children of mothers with university degrees scored significantly lower on externalising scores across models, while middle or higher household income was associated with significantly lower externalising and internalising scores across models. Polygenic scores for depression were associated with higher internalising and externalising symptoms, while polygenic scores for ADHD were associated with higher externalising scores. A comparison of model AIC scores showed that models which included polygenic scores were better supported than the models without polygenic scores (Supplementary Table ST2).

**Figure 1.**
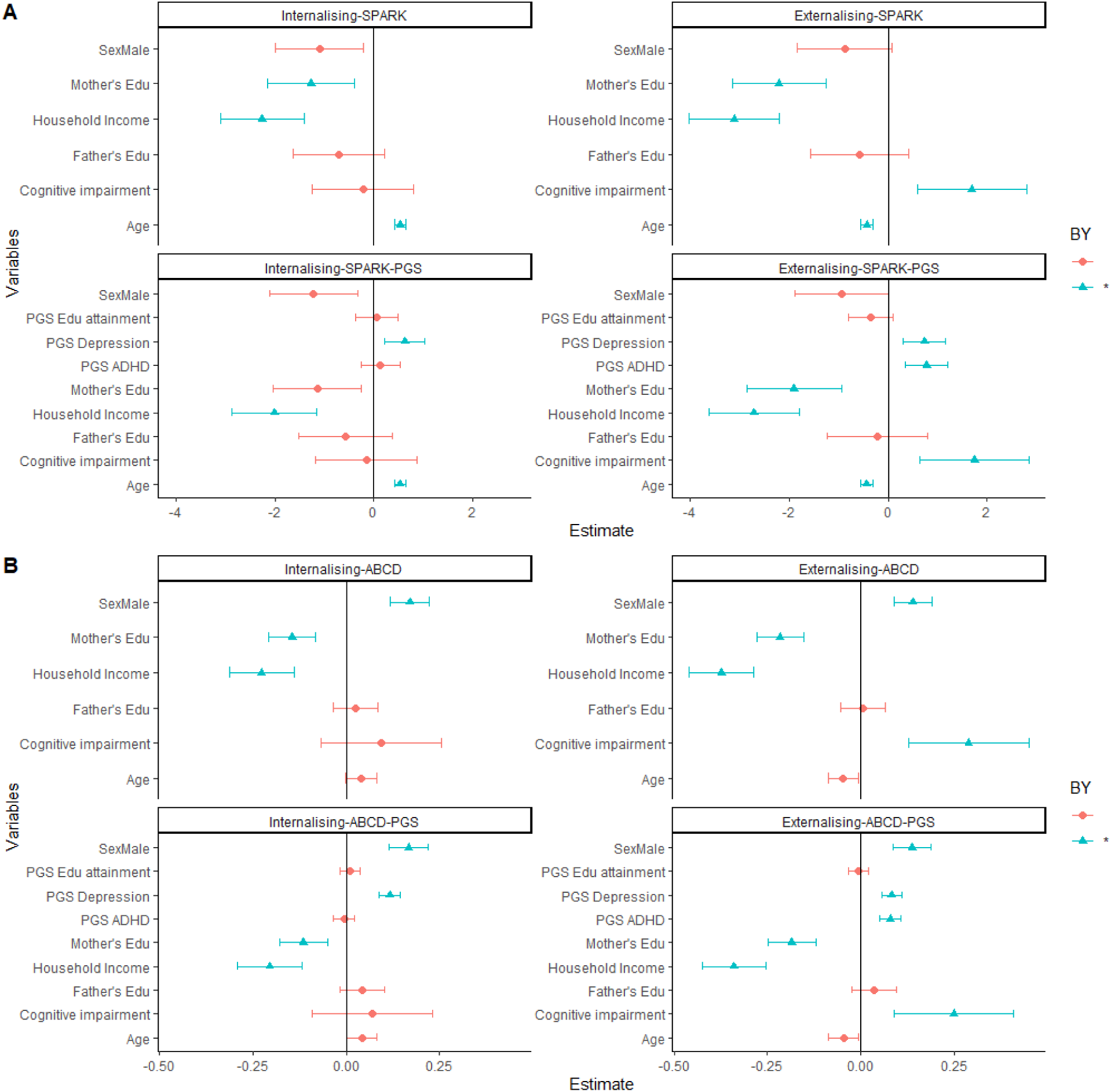
Regression models for (A) SPARK and (B) ABCD CBCL data, for models with and without PGS. Variables coloured green were significant after multiple testing correction. PGS: polygenic scores.

### Brain Phenotype Correlations

Given the modest genetic overlap between externalising symptoms in the autism and general population datasets, we next assessed whether the pattern of genetic correlations between different GWAS of externalising symptoms and structural and diffusion MRI phenotypes differed between the GWAS. We focussed on externalising symptoms as the low and non-significant SNP heritability of internalising symptoms GWAS made them unsuitable for genetic correlation analyses. Results of global brain correlations for externalising symptoms in SPARK, ABCD and Baselmans et al [21] are shown in Supplementary Table ST3, while regional correlations are shown in Supplementary Tables ST4-6. There was a consistent pattern of global and regional genetic correlations with brain phenotypes across datasets (Figure 2), with the strongest global associations being with Baselmans et al [21], which showed significant negative correlations after multiple testing correction with global measures of cortical expansion: folding index (rG=-1.27E-01, s.e.=3.15E-02, p_adj=4.99E-04), gray matter volume (rG=-1.46E-01, s.e.=2.94E-02, p_adj=2.31E-05), intrinsic curvature index (rG=-1.36E-01, s.e.=2.89E-02, p_adj=3.17E-05) and surface area (rG=-1.41E-01, s.e.=2.90E-02, p_adj=2.31E-05). Regional correlations for SPARK and ABCD externalising were largely in the same direction with the Baselmans et al (2021) externalising dataset (Figure 2, Supplementary Tables ST4 and ST5 and Supplementary Figure 2), but significant findings did not survive multiple testing correction. For the Baselmans et al (2021) externalising dataset, there were 289 significant correlations which survived FDR correction, spanning several brain regions, including negative correlations with gray matter volume, surface area, folding index and intrinsic curvature, as well as positive correlations with cortical thickness, mean diffusivity and isotropic volume fraction (Figure 3 and Supplementary Table ST6).

**Figure 2.**
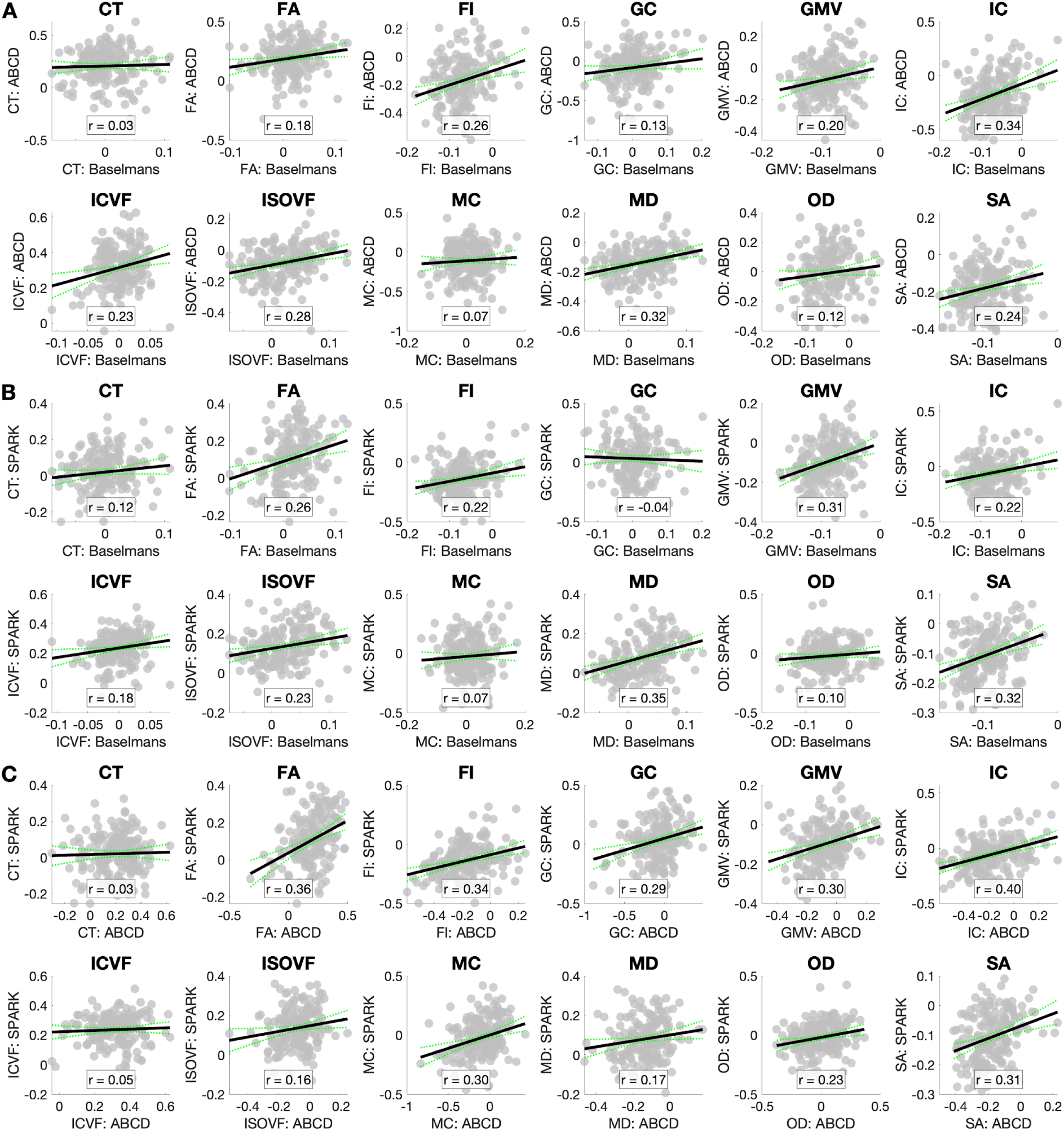
Scatterplot correlation of regional genetic correlations between the different externalising datasets. CT: cortical thickness; FA: fractional anisotropy; FI: folding index; GC: Gaussian curvature; GMV: gray matter volume; IC: intrinsic curvature; ICVF: intracellular volume fraction; ISOVF: isotropic volume fraction; MC: mean curvature; MD: mean diffusivity; OD: orientation diffusion index; SA: surface area.

**Figure 3.**
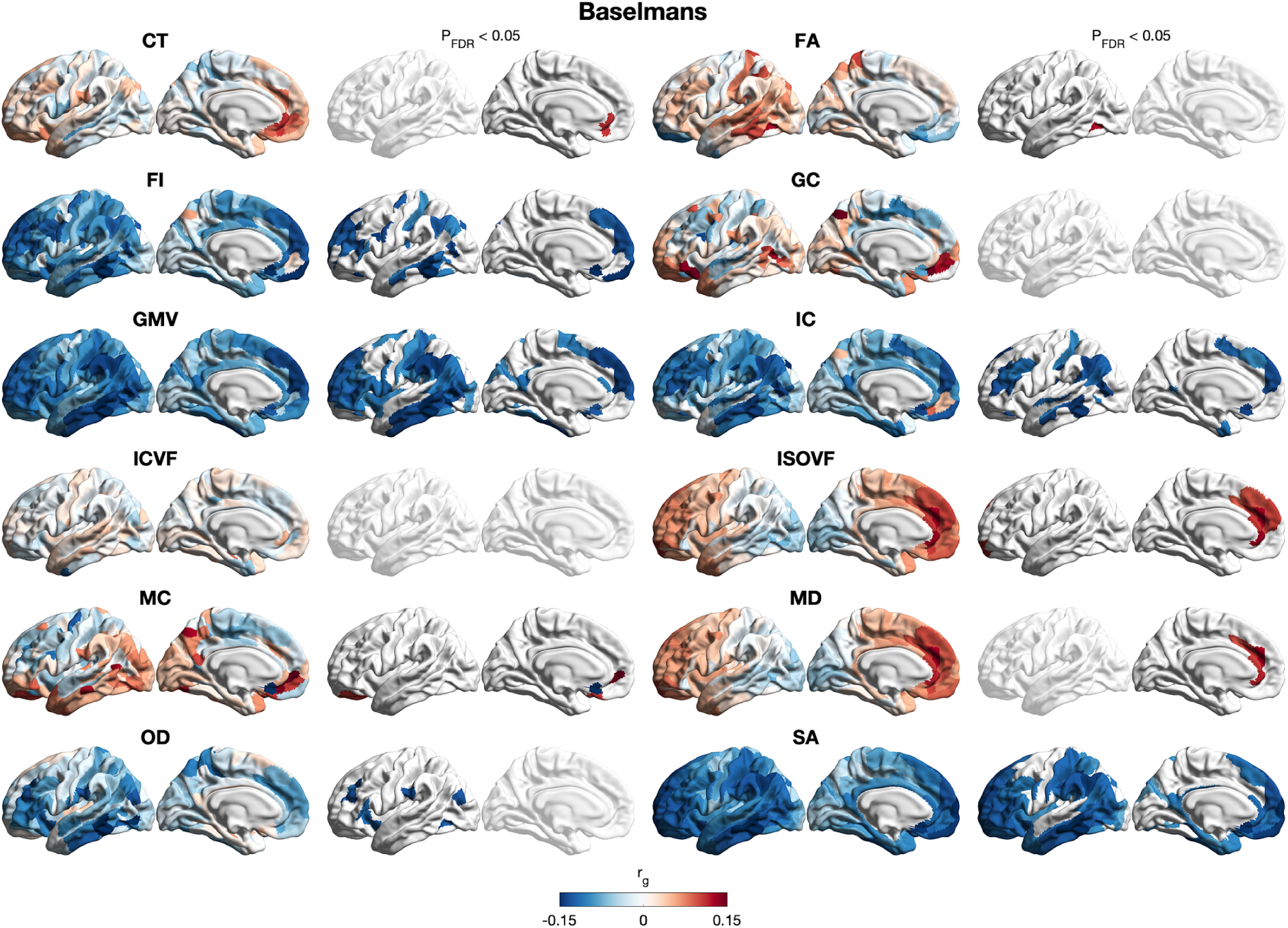
Plot of regional genetic correlations for externalising symptoms in the Baselmans et al [21] dataset, with each pair of brain surfaces representing medial and lateral views of the left hemisphere. CT: cortical thickness; FA: fractional anisotropy; FI: folding index; GC: Gaussian curvature; GMV: gray matter volume; IC: intrinsic curvature; ICVF: intracellular volume fraction; ISOVF: isotropic volume fraction; MC: mean curvature; MD: mean diffusivity; OD: orientation diffusion index; SA: surface area. Left panels: unthresholded genetic correlations. Right panels: genetic correlations at P_FDR_ < 0.05.

### Analysis of Cell Types

To investigate the specific brain cell types with gene enrichment associated with internalising and externalising symptoms across datasets, we investigated the enrichment in cell types of the adult human brain from Siletti et al [22] using MAGMA.

Results of cell-type enrichment analysis for the six datasets are shown in Supplementary Tables ST7-12. Only the best-powered dataset, Baselmans et al. [21], yielded cell-type enrichments that survived multiple-testing correction (Fig. 4a). For this dataset, all significant cell types were neurons. By dominant brain region, 22 mapped to the cerebral cortex, 7 to the hypothalamus, 7 to the amygdala, 3 to the midbrain, and 1 to the pons. Significant cell types were primarily GABAergic (inhibitory) and glutamatergic (excitatory). Notably, one of the top associations was a histaminergic (HDC⁺/VGLUT2⁺) hypothalamic cell type (#328), suggesting a potential role for histamine. Among the relatively independent significant cell types (n = 8), over-representation analyses localised signals to discrete dissections; Fig. 4b illustrates representative examples selected for being the top overrepresented dissection for each cell type: retrosplenial cortex (#132), dysgranular insular cortex (#241), dorsal anterior cingulate cortex (#283), lateral nucleus of the amygdala (#228), inferior colliculus (#355), and hypothalamus (#328, #331, #422). The most striking pattern was hypothalamic involvement: multiple enriched cell types were overrepresented in dissections from the mamillary, tuberal and supraoptic regions of the hypothalamus (Fig. 4b), a pattern less commonly highlighted in prior psychiatric cell-type analyses.

**Figure 4.**
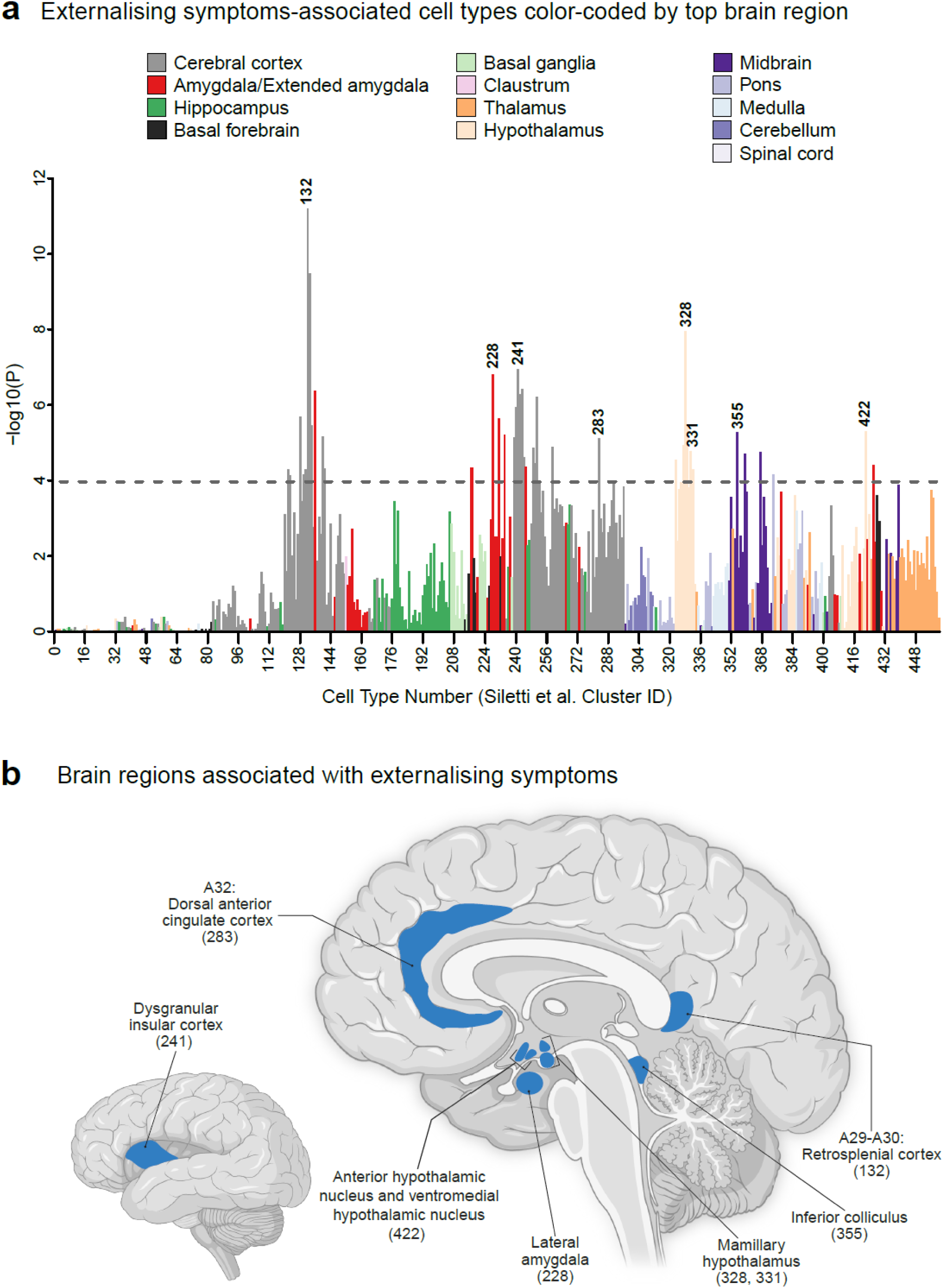
Cell type associations for externalising symptoms (Baselmans et al). a. Bar plot depicting associations with externalising symptoms across 461 cell types. Cell types are colour-coded by their dominant brain region. The x-axis shows cell-type numbers and the y-axis statistical significance (-log10(*p*)). The dashed horizontal line indicates the Bonferroni threshold for 461 tests (*p*<1.1×10^-4^). Bars corresponding to relatively independent significant cell types are labelled with their cell type numbers. b. Sagittal brain map highlighting brain regions that represent the top overrepresented dissections in relatively independent significant cell types. Labels indicate dissection names with corresponding cell type numbers (cluster IDs) in brackets.

### Colocalisation Analysis

Finally, to investigate whether internalising and externalising symptoms share causal loci with other phenotypes, we ran colocalisation analysis using *coloc* [23], which yielded results for the Baselmans et al [21] externalising dataset (Supplementary Table ST13). There were 38 configurations with PP.H4.abf>0.95, across 16 phenotypes (global brain cortical thickness, schizophrenia, bipolar disorder, ADHD anxiety, MDD, neuroticism, PTSD, substance use disorder, alcohol use disorder, daily cigarettes consumption, sleep duration, insomnia, BMI, educational attainment, and agreeableness).

## DISCUSSION

This study employed the tools of structural brain imaging and functional genomics to explore the features of internalising and externalising symptoms in autistic individuals (in the SPARK dataset) and individuals in general population datasets - ABCD, Baselmans et al [21] externalising, and Jami et al [20] internalising. The heritability estimates were similar for SPARK and ABCD. Genetic correlations were further calculated for the externalising and internalising datasets. Possibly due to low SNP heritability, there was no recorded genetic correlation estimate for the internalising datasets. There was however moderate correlation between ABCD and SPARK externalising, albeit with wide standard errors, likely due to limited statistical power. The genetic correlation between the general population cohorts was statistically higher than between the autism and general population, raising the question of whether externalising symptoms differ between autistic individuals and the general population. Possible explanations would include true genetic differences as well as possible differences in how externalising symptoms are observed or reported in autistic individuals and the general population. While Baselmans et al [21] had reported 50 GWAS-significant hits for the disruptive behaviour component of externalising symptoms in their much larger general population datasets, Jami et al [20] reported no GWAS-significant hits for internalising symptoms, albeit with a smaller sample than Baselmans et al [21]. There were no GWAS-significant hits for either externalising or internalising symptoms in SPARK or ABCD - expectedly, given the modest sample sizes in both datasets. There were however loci in the GWAS-suggestive range, with the possibility of reaching GWAS significance given larger sample sizes.

Among autistic individuals and the general population, regression models showed that presence of cognitive impairment among probands was associated with externalising symptoms. Male probands were also more likely to have both internalising and externalising symptoms in the general population but not in autistic population. In both autistic and general populations, higher maternal education was associated with lower externalising and internalising symptoms, supporting the role of environmental factors (in addition to genetics) in both symptom domains. This likely represents a complex gene-environment interplay, possibly indicative of genes that relate to high education of mothers as much as of environmental influence. Further work is required to parse the genetic (including direct and indirect effects) and environmental contribution. Higher household income was also associated with lower scores in both symptom domains in the autistic and general population. This being a cross-sectional study, it was not possible to infer causality; nevertheless, these findings highlight the need for integration of genetic and environmental factors in understanding the mechanisms underlying internalising and externalising symptoms - as previously observed [24], both genetic and environmental factors partially influence externalising symptoms, the same being likely true for internalising symptoms.

Among autistic individuals and the general population, there was also significant positive association between internalising symptoms and depression polygenic scores, and between externalising symptoms and both depression and ADHD polygenic scores. The finding of association between externalising symptoms and polygenic scores for depression may also reflect the considerable phenotypic overlap between internalising and externalising symptoms [2]. Similar associations between trajectories of internalising and externalising symptoms with polygenic scores for neuroticism were reported by Constantini et al [25], who observed that a genetic liability to neuroticism, measurable from early life through polygenic scores, was associated with severity and persistence of emotional and behavioural problems. While the current study is not longitudinal, there is a similar association between common genetic variation (as quantified by GWAS-derived polygenic scores) and the presence of internalising and externalising symptoms. With reference to broader genetic architecture, Wootton et al [26] found that both common and rare genetic variation contribute to internalising and externalising symptoms, with associations between externalising symptoms and polygenic indices for externalising behaviours, ADHD, and the cognitive component of educational attainment being primarily explained by direct genetic effects. For internalising symptoms, the evidence was less strong. No associations were found between either symptom domain and polygenic scores in the general population dataset.

Although there were promising brain imaging phenotype correlations for autistic individuals across symptom groups, and for internalising symptoms in the general population, the fact that these significant associations did not survive multiple testing correction makes it difficult to speculate on them. Aside from power limitations in the other datasets,differences in brain correlations could also be due to participant characteristics (such as age and sex) and the method of ascertainment (CBCL scores for SPARK and ABCD, composite measures for Baselmans et al [21]).

There was nevertheless a consistent direction of brain correlations across datasets, the strongest findings being with the best-powered Baselmans et al [21] dataset, which showed strong association with several measures of cortical expansion. Across gray matter volume, surface area, folding index, and intrinsic curvature, the strongest negative correlations in the Baselmans et al [21] dataset included regions of the dorsolateral and ventromedial prefrontal cortex, which are crucial for executive control, planning, and working memory (areas 9, 9–46d, 10d); orbitofrontal cortex and area 25 (subgenual ACC), involved in emotion regulation and reward valuation; the temporal cortex (involved in perceptual and associative processing); and the parietal-temporo-occipital junction, involved in integration and attention networks. Davis et al [16] reported that externalising symptoms were associated with greater gray matter volumes in the thalamus, caudate nuclei and the occipital pole, with lower volumes in the ventral striatum and amygdala. In addition, they found significant associations with intracellular volume fraction in the medial lemniscus, cerebral peduncle, and middle cerebellar peduncle. In a previous study, associations were found between coexisting (heterotypic) internalising and externalising symptom interactions and striatal, amygdalar, and anterior cingulate cortex volumes [27].

Beyond just identifying superclusters of interest, Fang et al [28] have demonstrated that transcriptomically defined cell clusters are derived from distinct cell layers. Reassuringly, the significantly enriched cell types in Baselmans et al [21] were from neuronal cells, as in previous analyses with mental health phenotypes. The majority of significant cell types were also of cortical origin; Davis et al [16] similarly reported that externalising genes were enriched in GABAergic, cortical and hippocampal neurons. The neuronal cell types associated with externalising symptoms were primarily from cortex, hypothalamus, and amygdala, with additional associated cell types from the midbrain and pons. Many of the neuronal cell types reported here overlap with cell types found to be associated with one or more psychiatric phenotypes examined previously, with three striking exceptions: hypothalamic cell types, histaminergic (HDC⁺) cell types, and a midbrain GABAergic cell type #355 with the strongest over-representation in the inferior colliculus and additional over-representation in the periaqueductal gray (PAG).

Seven of the forty externalising cell types were primarily from the hypothalamus, and these included three of the eight total relatively independent significant cell types. This latter finding of distinct hypothalamic cell types is particularly intriguing because it suggests the possibility that these cell types function together in a circuit, given that they are colocalized in the hypothalamus, and even within smaller subdivisions of the hypothalamus. Future work with spatial transcriptomics could be used to determine the spatial relationships among these cell types, providing further clues into their function and insight into brain circuits contributing to externalising symptoms.

Regarding other findings that are unexpected in that they are not overlapping with cell types previously reported to be associated with psychiatric disorders with this same method, we highlight that histaminergic (HDC⁺) neuronal cell types which were previously not associated with a psychiatric phenotype. Specifically, cell types #328 and #329 are both annotated as HDC⁺/VGLUT2⁺ hypothalamic neurons, and this suggests that histamine may play a role in externalising symptoms. The other “unexpected” cell type reported here is #355, an inhibitory (GABAergic) neuronal cell type with strongest enrichment in the inferior colliculus and additional enrichment in the PAG. The inferior colliculus is central to auditory processing, providing a possible link to reports of auditory-processing abnormalities in adolescent delinquency. One final cell type of interest, which was significant but not one of the relatively independent significant cell types, is pons cell type #374, an excitatory cell type found primarily in the parabrachial nucleus.

Overall, regarding externalising-associated cell types, we see a picture of substantially greater involvement of subcortical cell types – in the hypothalamus, midbrain, and the pons – than that seen with the other psychiatric disorders examined previously using the same methods (schizophrenia, bipolar disorder, and depression, being the best powered). Given the central role of the hypothalamus in arousal, and given experimental evidence of hypoarousal among individuals with high externalising symptoms, the hypothalamic cellular associations reported here may provide long-needed detail about the precise cellular basis of arousal differences among individuals with high externalising symptoms, including the specific cell types that might function together in specific microcircuits in the hypothalamus. Alternatively, these specific cell types might underlie or contribute to other aspects of externalising phenotypes. Nevertheless, it is useful to be able to parse polygenic risk for externalising symptoms into specific cellular contributions in the human brain, and we report here multiple highly specific human brain cell types linked to externalising symptoms. While there were promising observations with respect to autistic individuals, and for internalising symptoms in the general population, the fact that these did not survive multiple comparison means they have to be interpreted with caution pending more robust evidence in future studies.

*Coloc.abf* makes the assumption that only one variant is causal at a given locus [23]. While this is a simplification, it nevertheless enabled investigation of candidate shared causal variants between externalising symptoms and several phenotypes. The association with global cortical thickness, in conjunction with the finding of significant genetic correlation, provides strong evidence that externalising symptoms and global cortical thickness may not only be associated but might share causal genomic loci. Reassuringly, there were associations with typically considered externalising conditions such as schizophrenia, bipolar disorder and ADHD - this supports further investigation of externalising symptoms as a transdiagnostic corollary to these diagnostic categories. However, there were also associations with internalising conditions such as anxiety and depression, which serve as reminder that internalising and externalising symptoms are different but overlapping dimensions. Lastly, there were associations with several substance use phenotypes - Baselmans et al [21] similarly found evidence for bidirectional causal influences between the disruptive behaviour component of externalising symptoms and substance use behaviors.

One of the key strengths of this study is the analysis of cell types using what is currently the only comprehensive human brain cell atlas available [22]. The study was however limited by the relatively small size of all datasets except that of Baselmans et al [21], likely constraining the significance of GWAS findings as well as brain imaging and cell type correlations.

In conclusion, this study found that internalising and externalising symptoms are heritable, with stronger correlation between general population datasets than between autistic and general population. Among autistic individuals and the general population, internalising and externalising symptoms are associated with common polygenic variants as well as with environmental factors. Finally, the study provides strong evidence linking externalising symptoms in the general population with specific brain imaging phenotypes and cell types; the evidence for internalising symptoms (and for both symptom groups in autistic individuals) was less strong, possibly due to the power limitations of available datasets.

## METHODS

This study involved autistic individuals in the SPARK (2024-04-10) and ABCD datasets. Each dataset was subset to participants with available data on the Child Behaviour Checklist (CBCL). The CBCL[29] is available in two versions - a younger version for 1.5-to 5-year-olds, and an older version for 6- to 18-year-olds, and is a widely used tool for assessing internalising and externalising symptoms in children and adolescents, including those with an autism diagnosis [30]. This study was limited to the older (6-18 year old) version due to its sample size, and to enable comparison with ABCD for which data for the younger age group was not available. We did not meta-analyse the two versions due to concerns about the differences in reported factor structure. According to the Data Release Notes, individuals with more than 8 items missing were marked with a cbcl_validity_flag, scale scores of which may be invalid per publisher guidelines. All such individuals were removed from the analysis.

Regarding the other two datasets, Jami et al [20] included analysis of 64,561 children and adolescents between 3 and 18 years of age, across 22 cohorts, assessed for internalising symptoms. The authors performed multiple univariate GWASs and then aggregated the results in meta-analyses that accounted for sample overlap. Meta-analysis of overall internalising symptoms detected no genome-wide significant hits and showed low SNP heritability (1.66%, 95%CI = 0.84-2.48%, Neff=132,260). Baselmans et al [21] also performed multivariate genome-wide association meta-analyses using summary statistics on 13 externalising phenotypes with no specified age restriction. Genetic data was categorised into disruptive behaviour (DB) and risk taking behaviour (RTB) factors, with the statistics for DB, much more than RTB, signalling genetic predisposition to adverse cognitive, mental health, and personality outcomes. Genome-wide association meta-analyses for DB (Neff=523,150) yielded 50 independent signals, with SNP heritability of 0.0396 (SE=0.0015).

### Genomic Analysis

Quality control (QC) of genetic data in the SPARK and ABCD datasets followed standard procedures: the analysis was limited to individuals of European ancestry as provided by both datasets, with non-European ancestry individuals excluded due to power limitations. Post-imputation filters included a 92% SNP missingness threshold, 0.2 threshold for inbreeding coefficient and Hardy-Weinberg equilibrium (p < 1E-6). Genetic principal components were generated with PCAir in GENESIS[31.32], which accounts for relatedness among individuals, using a relatedness matrix generated using KING[33]. For the other two datasets, details of data and QC are available elsewhere [20,21].

SNP heritabilities and genetic correlations were calculated for SPARK and ABCD data using GCTA-GREML[34]. GWAS was done using fastGWAS[35] separately for SPARK (3763 individuals and 9,640,223 SNPs) and ABCD datasets (5678 individuals and 9,621,972 SNPs), with age at interview, informant-reported sex and the first 10 genetic principal components as covariates. Polygenic scores for depression, ADHD and educational attainment were calculated using PRScs [36], using publicly available summary statistics from the most recent GWAS for ADHD [37], depression [38] and educational attainment [39].

### Regression Modelling

Multiple logistic regression analysis within the generalised linear model was conducted for individuals included in the SPARK and ABCD datasets. Models were computed separately with CBCL internalising and externalising scores as dependent variables. The first set of models included age, sex, presence/absence of cognitive impairment, as well as mother’s and father’s highest education, and annual household income. The second set of models had, in addition, polygenic scores for ADHD, depression and educational attainment, as well as the first ten genetic principal components, as covariates. Informer-reported professional diagnoses of cognitive impairment were available in SPARK. For ABCD, we converted the standardised total composite cognitive scores (which are adjusted for age and sex) to the IQ scale, and categorised scores falling below two standard deviations from the mean (IQ<70) as cognitive impairment. The highest educational level attained by each biological parent was coded as “university_degree” or “high_school_or_other”, and annual total household income was coded as “lower” (50,000 USD or less) or “middle_or_higher” (51,000 or higher) in both datasets. Multiple testing correction was done using the Benjamini-Yekutieli method of false discovery rate.

### Brain Phenotype Correlations

For all datasets except Jami et al [20], genetic correlations were calculated separately with 12 structural global brain phenotypes (cortical thickness, fractional anisotropy, folding index, Gaussian curvature, gray matter volume, intrinsic curvature index, intracellular volume fraction, isotropic volume fraction, mean curvature, mean diffusivity, orientation diffusion index and surface area), using summary statistics from Warrier et al [9]. The summary statistics were averaged across the hemispheres, hence 180 regions per phenotype, and the AVI region was missing for the folding index (hence 2,159 instead of 2,160 regions in total). Regions were based on Glasser (or HCP-MMP1.0) parcellations[40]. The genetic correlations were calculated using LDSC [41,42], with FDR correction.

### Analysis of Cell Types

Cell type associations with externalising and internalising symptoms across six datasets – SPARK (externalising and internalising), ABCD (externalising and internalising), Baselmans et al. [21], and Jami et al. [20] – were examined using MAGMA (v1.10)[43], following the methodology of Duncan et al. [44]. Human brain single-nucleus RNA sequencing data were obtained from Siletti et al. [22], comprising 3,369,219 nuclei from 107 dissections grouped into 461 transcriptomic cell types. MAGMA was first applied to compute gene-level *p*-values from GWAS summary statistics and then used to test whether trait-associated genes were preferentially expressed in specific brain cell types based on expression-specificity scores. A conservative Bonferroni correction was used to define significant associations: *p*<0.05/461 ≈ 1.1×10^-4^, and conditional analyses were conducted to identify relatively independent signals among all significant cell types.

For the anatomical panel (Fig. 4b), to aid interpretation, we depicted the significant, relatively independent cell types and assigned each to a single representative brain dissection, although cell types were found in multiple dissections in the Siletti atlas. To select the brain region to depict, we tested overrepresentation of each cell type across dissections using Fisher’s exact test against a pooled background of all cell types, applied Benjamini–Hochberg false discovery rate (FDR) control within each cell type, and ranked dissections by observed/expected (O/E) enrichment, with expected counts computed from the pooled background proportion of cells in each dissection. We shaded the top overrepresented dissection per cell type from the significant, enriched set (q<0.05; O/E>1). For a complete list of brain dissections contributing cells to each cell type, see Supplementary Table ST7-12.

### Colocalisation

We performed colocalisation analysis using *coloc* [23]. Of all the datasets - SPARK, ABCD, Jami et al [20] and Baselmans et al [21] - only Baselmans et al [21] showed significant clumping, so analysis was limited to this dataset, which had 40 unique LD blocks. For each sentinel SNP in the clumped output, we extracted the corresponding LD block and performed *coloc* runs for the 48 phenotypes (covering psychiatric, neurologic, cognitive, personality and other traits, thus giving 1,920 potential runs) using coloc.abf, which estimates the posterior probabilities of different causal variant configurations under the assumption of a single causal variant per trait.

## ACKNOWLEDGEMENTS

SBC received funding from the Wellcome Trust 214322\Z\18\Z. For the purpose of Open Access, the author has applied a CC BY public copyright licence to any Author Accepted Manuscript version arising from this submission. SBC also received funding from the Innovative Medicines Initiative 2 Joint Undertaking under grant agreement No 777394 for the project AIMS-2-TRIALS. This Joint Undertaking receives support from the European Union’s Horizon 2020 research and innovation programme and EFPIA and AUTISM SPEAKS, Autistica, SFARI. SBC also received funding from the Autism Centre of Excellence (now Autism Action), SFARI, the Templeton World Charitable Fund and the MRC. We are grateful to Cambridge University Development and Alumni Relations (CUDAR) for anonymous donations. The funders had no role in the design of the study; in the collection, analyses, or interpretation of data; in the writing of the manuscript, or in the decision to publish the results. Any views expressed are those of the author(s) and not necessarily those of the funders (including IHI-JU2).

## ETHICS STATEMENT

The Cambridge University Human Biology Research Ethics Committee provided ethical approval to access and analyse de-identified data from SPARK and ABCD. The SPARK study is funded by the Simons Foundation and uses a single, central Institutional Review Board (WCG IRB Protocol #20151664). Informed consent was obtained by the SPARK study team from the participant or the caregiver/parent. Ethical approval for ABCD and other datasets was obtained from multiple institutional review boards.

## REPORTING SUMMARY

## DATA AVAILABILITY

SPARK data were accessed via SFARI Base and ABCD data via the NIMH Data Archive under controlled access. GWAS summary statistics from Jami et al. (2022) are publicly available from the GWAS Catalog (EFO_0020971). Summary statistics from Baselmans et al. (2021) were obtained from the authors and are not publicly available. No new individual-level data were generated.

## CODE AVAILABILITY

Bespoke code for heritability and genetic correlation estimates, GWAS and regression modelling is available on Github (https://github.com/NiranOkewole/CamPhD2).

The code used for our implementation of the cell type association analysis is available on GitHub (https://github.com/Integrative-Mental-Health-Lab/linking_cell_types_to_brain_phenotypes).

---

**Supplementary Figure 1.**
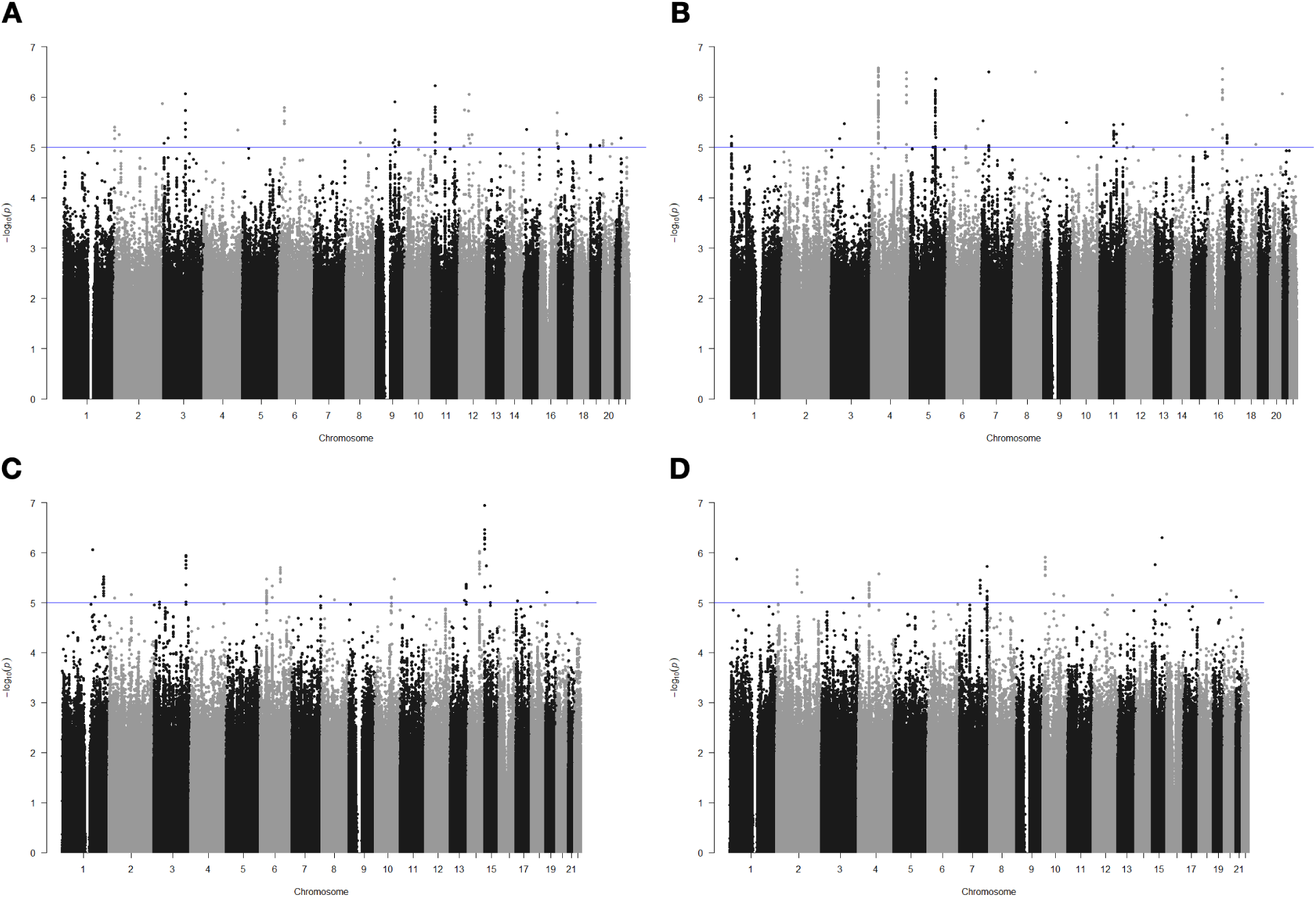
Manhattan plots of internalising and externalising symptoms in SPARK and ABCD. A: SPARK internalising B: SPARK externalising. C: ABCD internalising D: ABCD externalising. The blue line represents p value < 5e-5, conventionally taken as in the suggestive range for GWAS.

**Supplementary Figure 2.**
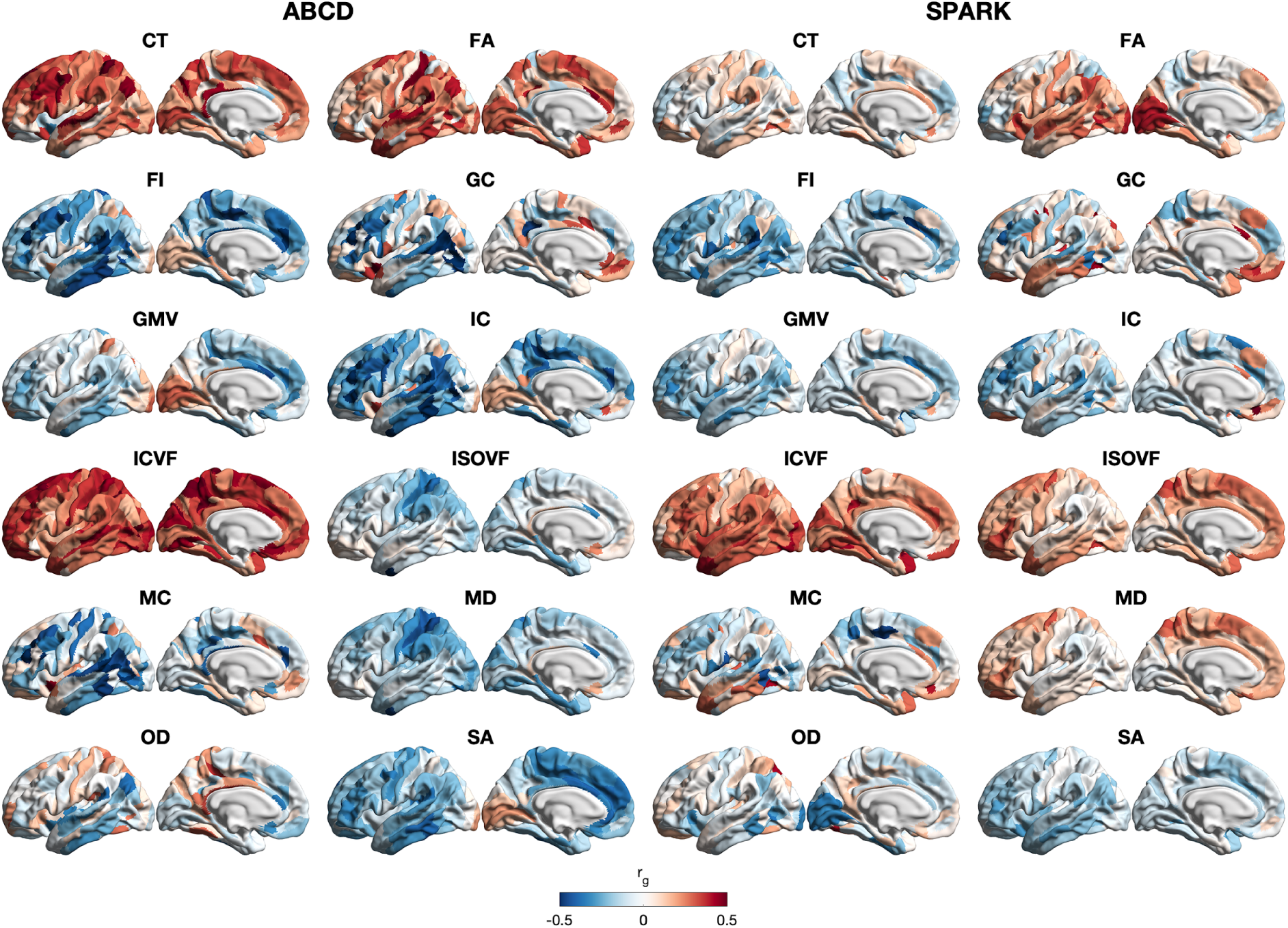
Unthresholded regional genetic correlations for ABCD and SPARK. CT: cortical thickness; FA: fractional anisotropy; FI: folding index; GC: Gaussian curvature; GMV: gray matter volume; IC: intrinsic curvature; ICVF: intracellular volume fraction; ISOVF: isotropic volume fraction; MC: mean curvature; MD: mean diffusivity; OD: orientation diffusion index; SA: surface area.

## REFERENCES

1. Astle, D. E. et al. Annual Research Review: The transdiagnostic revolution in neurodevelopmental disorders. J. Child Psychol. Psychiatry 63, 397–417 (2022). 10.1111/jcpp.13481

2. Achenbach, T. M., et al. Internalizing/Externalizing Problems: Review and Recommendations for Clinical and Research Applications. J. Am. Acad. Child Adolesc. Psychiatry 55, 647–656 (2016). 10.1016/j.jaac.2016.05.012

3. Grotzinger, A.D. et al. Mapping the genetic landscape across 14 psychiatric disorders. Nature, 10.1038/s41586-025-09820-3. 10 Dec. 2025, doi:10.1038/s41586-025-09820-3

4. Blanken, L. M. et al. Cognitive functioning in children with internalising, externalising and dysregulation problems: a population-based study. Eur. Child Adolesc. Psychiatry 26, 445–456 (2017). 10.1007/s00787-016-0903-9

5. Nakua, H. et al. Investigating cross-sectional and longitudinal relationships between brain structure and distinct dimensions of externalizing psychopathology in the ABCD sample. Neuropsychopharmacology 50, 499–506 (2025). 10.1038/s41386-024-02000-3

6. Antaki, D. et al. A phenotypic spectrum of autism is attributable to the combined effects of rare variants, polygenic risk and sex. Nat. Genet. 54, 1284–1292 (2022). 10.1038/s41588-022-01064-5

7. Warrier, V. et al. Genetic correlates of phenotypic heterogeneity in autism. Nat. Genet. 54, 1293–1304 (2022). 10.1038/s41588-022-01072-5

8. Shen, X. et al. A phenome-wide association and Mendelian Randomisation study of polygenic risk for depression in UK Biobank. Nat. Commun. 11, 2301 (2020). 10.1038/s41467-020-16022-0

9. Warrier, V. et al. Genetic insights into human cortical organization and development through genome-wide analyses of 2,347 neuroimaging phenotypes. Nat. Genet. 55, 1483–1493 (2023). 10.1038/s41588-023-01475-y

10. Stauffer, E. M. et al. The genetic relationships between brain structure and schizophrenia. Nat. Commun. 14, 7820 (2023). 10.1038/s41467-023-43567-7

11. Gu, Y. et al. Polygenic scores for autism are associated with reduced neurite density in adults and children from the general population. Mol. Psychiatry 30, 3393–3403 (2025). 10.1038/s41380-025-02927-z

12. Whittle, S., Vijayakumar, N., Simmons, J. G. & Allen, N. B. Internalizing and Externalizing Symptoms Are Associated With Different Trajectories of Cortical Development During Late Childhood. J. Am. Acad. Child Adolesc. Psychiatry 59, 177–185 (2020). 10.1016/j.jaac.2019.04.006

13. Jarvers, I. et al. The relationship between adolescents’ externalizing and internalizing symptoms and brain development over a period of three years. NeuroImage Clin. 36, 103195 (2022). 10.1016/j.nicl.2022.103195

14. Yao, S. et al. Connecting genomic results for psychiatric disorders to human brain cell types and regions reveals convergence with functional connectivity. Nature communications vol. 16,1 395. 4 Jan. 2025, doi:10.1038/s41467-024-55611-1

15. Davis, C. N. et al. Integrating HiTOP and RDoC frameworks Part I: Genetic architecture of externalizing and internalizing psychopathology. Psychol. Med. 55, e138 (2025). doi:10.1017/S0033291725000856

16. Davis, C. N. et al. Integrating HiTOP and RDoC frameworks part II: shared and distinct biological mechanisms of externalizing and internalizing psychopathology. Psychol. Med. 55, e137 (2025). 10.1017/S0033291725000819

17. Kotov, R. et al. The Hierarchical Taxonomy of Psychopathology (HiTOP): A dimensional alternative to traditional nosologies. J. Abnorm. Psychol. 126, 454–477 (2017). 10.1037/abn0000258

18. Insel, T. et al. Research domain criteria (RDoC): toward a new classification framework for research on mental disorders. Am. J. Psychiatry 167, 748–751 (2010). 10.1176/appi.ajp.2010.09091379

19. Liang, Y. et al. BrainXcan identifies brain features associated with behavioral and psychiatric traits using large-scale genetic and imaging data. Dev. Cogn. Neurosci. 73, 101542 (2025). 10.1016/j.dcn.2025.101542

20. Jami, E. S. et al. Genome-wide Association Meta-analysis of Childhood and Adolescent Internalizing Symptoms. J. Am. Acad. Child Adolesc. Psychiatry 61, 934–945 (2022). doi:10.1016/j.jaac.2021.11.035

21. Baselmans, B. et al. The Genetic and Neural Substrates of Externalizing Behavior. Biol. Psychiatry Glob. Open Sci. 2, 389–399 (2021). 10.1016/j.bpsgos.2021.09.007

22. Siletti, K. et al. Transcriptomic diversity of cell types across the adult human brain. Science 382, eadd7046 (2023). doi:10.1126/science.add7046

23. Wallace, C. A more accurate method for colocalisation analysis allowing for multiple causal variants. PLoS Genet. 17, e1009440 (2021). 10.1371/journal.pgen.1009440

24. Newsome, J., Boisvert, D. & Wright, J. P. Genetic and environmental influences on the co-occurrence of early academic achievement and externalizing behavior. J. Crim. Justice 42, 45–53 (2014). https://psycnet.apa.org/doi/10.1016/j.jcrimjus.2013.12.002

25. Costantini, I. et al. Childhood trajectories of internalising and externalising problems associated with a polygenic risk score for neuroticism in a UK birth cohort study. JCPP Adv. 3, e12141 (2023). doi:10.1002/jcv2.12141

26. Wootton, O. et al. The Contribution of Common and Rare Genetic Variation to Emotional and Behavioural Symptoms in Childhood and Adolescence. medRxiv 2025.07.01.25330628 (2025). 10.1101/2025.07.01.25330628

27. Schettini, E., Wilson, S. & Beauchaine, T. P. Internalizing-externalizing comorbidity and regional brain volumes in the ABCD study. Dev. Psychopathol. 33, 1620–1633 (2021). doi:10.1017/s0954579421000560

28. Fang, R. et al. Conservation and divergence of cortical cell organization in human and mouse revealed by MERFISH. Science 377, 56–62 (2022). doi:10.1126/science.abm1741

29. Achenbach, T. M. & Ruffle, T. M. The Child Behavior Checklist and related forms for assessing behavioral/emotional problems and competencies. Pediatr. Rev. 21, 265–271 (2000). 10.1542/pir.21-8-265

30. Pandolfi, V., Magyar, C. I. & Dill, C. A. An Initial Psychometric Evaluation of the CBCL 6-18 in a Sample of Youth with Autism Spectrum Disorders. Res. Autism Spectr. Disord. 6, 96–108 (2012). 10.1016/j.rasd.2011.03.009

31. Gogarten, S. M. et al. Genetic association testing using the GENESIS R/Bioconductor package. Bioinformatics 35, 5346–5348 (2019). 10.1093/bioinformatics/btz567

32. Conomos, M. P., Miller, M. B. & Thornton, T. A. Robust inference of population structure for ancestry prediction and correction of stratification in the presence of relatedness. Genet. Epidemiol. 39, 276–293 (2015). 10.1002/jcv2.12141

33. Manichaikul, A. et al. Robust relationship inference in genome-wide association studies. Bioinformatics 26, 2867–2873 (2010). 10.1093/bioinformatics/btq559

34. Yang, J., Lee, S. H., Goddard, M. E. & Visscher, P. M. GCTA: a tool for genome-wide complex trait analysis. Am. J. Hum. Genet. 88, 76–82 (2011). 10.1016/j.ajhg.2010.11.011

35. Jiang, L., Zheng, Z., Fang, H. & Yang, J. A generalized linear mixed model association tool for biobank-scale data. Nat. Genet. 53, 1616–1621 (2021). 10.1038/s41588-021-00954-4

36. Ge, T., Chen, C. Y., Ni, Y., Feng, Y. A. & Smoller, J. W. Polygenic prediction via Bayesian regression and continuous shrinkage priors. Nat. Commun. 10, 1776 (2019). 10.1038/s41467-019-09718-5

37. Demontis, D. et al. Genome-wide analyses of ADHD identify 27 risk loci, refine the genetic architecture and implicate several cognitive domains. Nat. Genet. 55, 198–208 (2023). 10.1038/s41588-022-01285-8

38. Als, T. D. et al. Depression pathophysiology, risk prediction of recurrence and comorbid psychiatric disorders using genome-wide analyses. Nat. Med. 29, 1832–1844 (2023). 10.1038/s41591-023-02352-1

39. Okbay, A. et al. Polygenic prediction of educational attainment within and between families from genome-wide association analyses in 3 million individuals. Nat. Genet. 54, 437–449 (2022). 10.1038/s41588-022-01016-z

40. Glasser, M. F. et al. A multi-modal parcellation of human cerebral cortex. Nature 536, 171–178 (2016). 10.1038/nature18933

41. Bulik-Sullivan, B. K. et al. LD Score regression distinguishes confounding from polygenicity in genome-wide association studies. Nat. Genet. 47, 291–295 (2015). 10.1038/ng.3211

42. Bulik-Sullivan, B. et al. An atlas of genetic correlations across human diseases and traits. Nat. Genet. 47, 1236–1241 (2015). 10.1038/ng.3406

43. de Leeuw, C. A., Mooij, J. M., Heskes, T. & Posthuma, D. MAGMA: generalized gene-set analysis of GWAS data. PLoS Comput. Biol. 11, e1004219 (2015). 10.1371/journal.pcbi.1004219

44. Duncan, L. E. et al. Mapping the cellular etiology of schizophrenia and complex brain phenotypes. Nat. Neurosci. 28, 248–258 (2025). doi:10.1038/s41593-024-01834-w

